# An archetypes approach to malaria intervention impact mapping: a new framework and example application

**DOI:** 10.1101/2022.08.01.22278276

**Authors:** Amelia Bertozzi-Villa, Caitlin Bever, Jaline Gerardin, Joshua L. Proctor, Meikang Wu, Dennis Harding, T. Deirdre Hollingsworth, Samir Bhatt, Peter W. Gething

## Abstract

**Background:** As both mechanistic and geospatial malaria modeling methods become more integrated into malaria policy decisions, there is increasing demand for strategies that combine these two methods. This paper introduces a novel archetypes-based methodology for generating high-resolution intervention impact maps based on mechanistic model simulations. An example configuration of the framework is described and explored.

**Methods:** First, dimensionality reduction and clustering techniques were applied to rasterized geospatial environmental and mosquito covariates to find archetypal malaria transmission patterns. Next, mechanistic models were run on a representative site from each archetype to assess intervention impact. Finally, these mechanistic results were reprojected onto each pixel to generate full maps of intervention impact. The example configuration used ERA5 and Malaria Atlas Project covariates, singular value decomposition, k-means clustering, and the Institute for Disease Modeling’s EMOD model to explore a range of three-year malaria interventions primarily focused on vector control and case management.

**Results:** Rainfall, temperature, and mosquito abundance layers were clustered into ten transmission archetypes with distinct properties. Example intervention impact curves and maps highlighted archetype-specific variation in efficacy of vector control interventions. A sensitivity analysis showed that the procedure for selecting representative sites to simulate worked well in all but one archetype.

**Conclusion:** This paper introduces a novel methodology which combines the richness of spatiotemporal mapping with the rigor of mechanistic modeling to create a multi-purpose infrastructure for answering a broad range of important questions in the malaria policy space. It is flexible and adaptable to a range of input covariates, mechanistic models, and mapping strategies and can be adapted to the modelers’ setting of choice.

## Background

Malaria is one of humanity’s oldest and most insidious ailments, co-evolving with mosquitoes and humans over millions of years [1]. This long history, combined with its complex transmission pathways, makes malaria a uniquely heterogeneous and environmentally sensitive disease. Understanding malaria in a given location requires not just an understanding of human behavior, movement, and demographics, but also a detailed knowledge of resident mosquito species and their behavior, local climate and hydrology, and the seasonal patterns of the landscape.

Computational modeling and malaria policy have been linked since the 1950s, from seminal pen-and-paper equations by Ronald Ross and George Macdonald to today’s detailed computational models capable of simulating individual humans, mosquitoes, climates, immune responses, malaria interventions, and much more. The fundamental goal of these models is to help decision-makers craft a malaria strategy that aligns with the epidemiological, economic, and cultural circumstances at hand.

The two main classes of malaria model in use today are mechanistic transmission models, which explicitly simulate disease spread in a population, and spatiotemporal models, which utilize geospatially-referenced covariates and classical statistical methods to generate estimated maps of malaria burden, intervention coverage, and other malaria-relevant features. The Malaria Atlas Project (MAP) is known for its high-resolution maps of malaria-related variables [2, 3, 4, 5, 6, 7, 8, 9, 10]. Over the last 15 years a number of mechanistic modeling groups, most prolifically at Imperial College, the Swiss Tropical and Public Health Institute, Northwestern University, PATH, the Ifakara Health Institue, the MORU Tropical Health Network, and the Institute for Disease Modeling (IDM), have supported decision-making at the local, national, and global scale [11, 12, 13, 14, 15].

Mechanistic and spatiotemporal models serve complementary purposes in malaria. Spatiotemporal models elucidate the past and present of disease burden and related metrics, which mechanistic models can then use for insight into the future. Maps provide a rich descriptive landscape, while mechanistic models contribute causal structure and exploration of counterfactual or hypothetical scenarios. As both modeling methods have grown in popularity and demand, there is increased interest in products that combine the two approaches, allowing for a spatially and temporally detailed exploration of the consequences of different policy decisions. This paper introduces a novel methodology which combines the richness of spatiotemporal mapping with the rigor of mechanistic modeling to create a multi-purpose infrastructure for answering a broad range of important questions in the malaria policy space.

Mechanistic models draw strength from their ability to replicate detailed relationships, but suffer several limitations. First, these detailed models require a large amount of input data to configure and calibrate to a particular setting. When tasked with simulating settings without such a rich data space, mechanistic models can be a challenge to appropriately configure. Second, mechanistic model complexity carries with it an associated computational cost. A single simulation can take minutes or hours to run, even on high-performance computing systems, often limiting the number of scenarios or locations that can reasonably be simulated. In contrast to these challenges, spatiotemporal methods are designed with incomplete data as an assumption and perform well at high resolutions [16, 17]. However, these statistical methods crucially lack the explicit causal relationships that allow mechanistic models to effectively test the consequences of different policies.

In the past, mechanistic modeling analyses working at the continental scale in Africa have utilized a range of methods to address these practical and computational challenges. Griffin et al. [18] modeled a range of stylized seasonality patterns and mosquito species mixes to assess intervention strategies. While this approach showcases hypothetical scenarios, there is no explicit link between any true location and any stylized seasonal profile, making geographic trends or burden estimation impossible. Walker et al. [11] grouped rainfall, entomological, and transmission intensity spatial covariates into three, four, and 18 clusters respectively, and ran simulations on all possible combinations of these groups before re-projecting onto the pixel level. This exhaustive strategy does not take advantage of the spatial relationships between different covariates, which could reduce computational burden and generate more informative cluster properties.

The methodology presented in this paper leverages the strength of both mechanistic and spatiotemporal methods to allow computationally-feasible generation of high-resolution maps that reflect mechanistically modeled scenarios. This process generates a range of archetypal seasonal and entomological profiles that can be useful for exploratory data analysis while also generating an explicit link between these profiles and any given location across Africa. This mapping capacity provides a computationally efficient pathway for presenting results geospatially and in terms of expected burden change.

Here, high-resolution, high-dimensional spatial covariates and machine learning methods are harnessed to generate a small number of spatially-explicit “archetypes” of malaria transmission, characterized by their covariate similarity. Next, mechanistic models are run on a representative site from each archetype, rather than on every pixel individually. Finally, spatial data and model results create a lookup table through which maps of intervention impact are generated.

Sorting many observations of high-dimensional data into groups by similarity is a common problem in machine learning. A common solution is to deploy a two-step process of first reducing the dimensionality of the dataset, and then clustering this reduced-dimensional data into groups [19, 20, 21]. This approach is appealing for its flexibility, but requires a number of specific decisions that can materially impact results. Namely:

1. Which covariates should be selected, and how should they be standardized?
2. Which algorithm should be used for dimensionality reduction?
3. Which algorithm should be used for clustering, and how should the number of clusters be determined?

When clustered results are being used as inputs for mechanistic models and subsequent generation of new maps, additional questions emerge:

4 How should representative sites be selected from each cluster?
5 Which mechanistic model should be used to assess intervention impact?
6 How should results from representative sites be reprojected back onto other members of the group?

This paper reviews each of these questions in turn, beginning with a discussion of the choices available in each space and their implications. As a demonstration, it describes the results of an eradication feasibility exercise undertaken by IDM and MAP in 2018. The original work was conducted by request of international stakeholders seeking guidance on whether eradication might be possible under highly optimistic environmental circumstances and malaria control scenarios. It harnessed spatiotemporal climate and mosquito covariates, IDM’s EMOD malaria transmission model, and MAP’s malaria prevalence maps to generate scenarios of malaria intervention impact at the 5km-by-5km pixel level for all of sub-Saharan Africa in 2050. The original analysis [22, 23] utilized an earlier version of the framework, and the version presented here represents a validation check to ensure that similar results arise from a more in-depth approach. A sensitivity analysis exploring the representativeness of a given “representative site” is also described.

The primary goal of this document is to introduce a novel modeling paradigm in sufficient detail to be adapted for other use cases. Throughout, the terms “archetypes strategy” and “archetypes framework” will be used to describe the general methodology of using dimensionality reduction and clustering to locate a subset of sites appropriate for mechanistic modeling. The term “example configuration” will be used to describe the specific set of choices within the broader strategy that are shown here for demonstration. The archetypes strategy is flexible and generalizable to any covariate set, mechanistic modeling platform, and geographic scale. The methods sections are subdivided into a general discussion and an explicit description of the example configuration parameters. The results section focuses on describing example configuration outcomes. Finally, the discussion section highlights lessons learned and other use cases of this archetypes framework.

## Methods

In the sections below, each of the six methodological questions described in the introduction is explored in detail. General theory is described first, followed by a description of the specific choices made in the example configuration. For the example configuration, simplicity was favored, but more complex options are noted.

### Covariate Selection

#### General

The covariates used for clustering should be selected mindfully to capture the types of variation to which the transmission model is most sensitive. For malaria, these should almost always include covariates that capture the different seasonality patterns of different malaria-endemic regions, as this directly impacts disease seasonality. For models that explicitly simulate different mosquito behaviors by species, covariates describing relative species abundance are also valuable. Baseline malaria transmission intensity is another useful source of input data, but its inclusion depends on the use case. The example configuration described below intentionally excluded it from the clustering process, instead running simulations for each archetype over a range of transmission intensities. Other possible covariates of interest include intervention history, health care accessibility, population demographics, and other behavioral inputs. Covariate data must geographically cover the region of interest, and should be utilized on the same spatial resolution as the final results. Covariate data for malaria will often also include a temporal component showing either seasonality or secular time.

Covariates are collected into a “stack” of spatial or spatiotemporal input files measuring different metrics on different scales. It is important to normalize or rescale these covariates prior to performing dimensionality reduction to avoid an artificial effect due to differently-scaled inputs. Even after normalization, however, a decision must be made regarding the relative weight of different inputs. For instance, in the example configuration below, clustering covariates included 12 rainfall layers, 12 temperature layers, and three mosquito species relative abundance layers. These covariates were not differentially weighted prior to dimensionality reduction, meaning that the rainfall and temperature layers had a stronger impact on results than the mosquito abundance layers. This choice was acceptable for the work at hand, but different circumstances might encourage different weighting.

#### Example Configuration

Environmental covariate data on rainfall and air temperature were sourced from the ERA5 project (https://www.ecmwf.int/en/forecasts/datasets/reanalysis-datasets/era5), a global reanalysis that generates internally-consistent estimates for a wide range of climate parameters across the globe. Specifically, monthly mean total precipitation and 2-meter air temperature from 2000 to 2018 were downloaded. Monthly means were averaged across the time series to generate synoptic 12-month trends. Because ERA5 spatial resolution is 0.25°-by-0.25°(approximately 30km-by-30km at the equator), covariates were resampled down to the 5km-by-5km level and realigned to match MAP’s standard spatial specifications.

Covariate data on the relative abundance of *Anopheles arabiensis, funestus*, and *gambiae* were obtained from Sinka et al. [6]. These values are static estimates and did not require resampling as they already met MAP spatial specifications.

All covariate rasters were masked to align with MAP estimates of *Plasmodium falciparum* transmission limits, and pixel-level values extracted into a nonspatial dataset for further analysis. Once extracted, pixel-level values of air temperature and rainfall were rescaled to fall between zero and one. Because they contained some extreme values that severely skewed the distribution, the most extreme 1% of rainfall values were reassigned to the 99th percentile value before being rescaled. Relative vector abundance values are proportions, and therefore these values already fell between zero and one and did not require rescaling. See Figure 1 for covariate distributions before and after rescaling.

**Figure 1.**
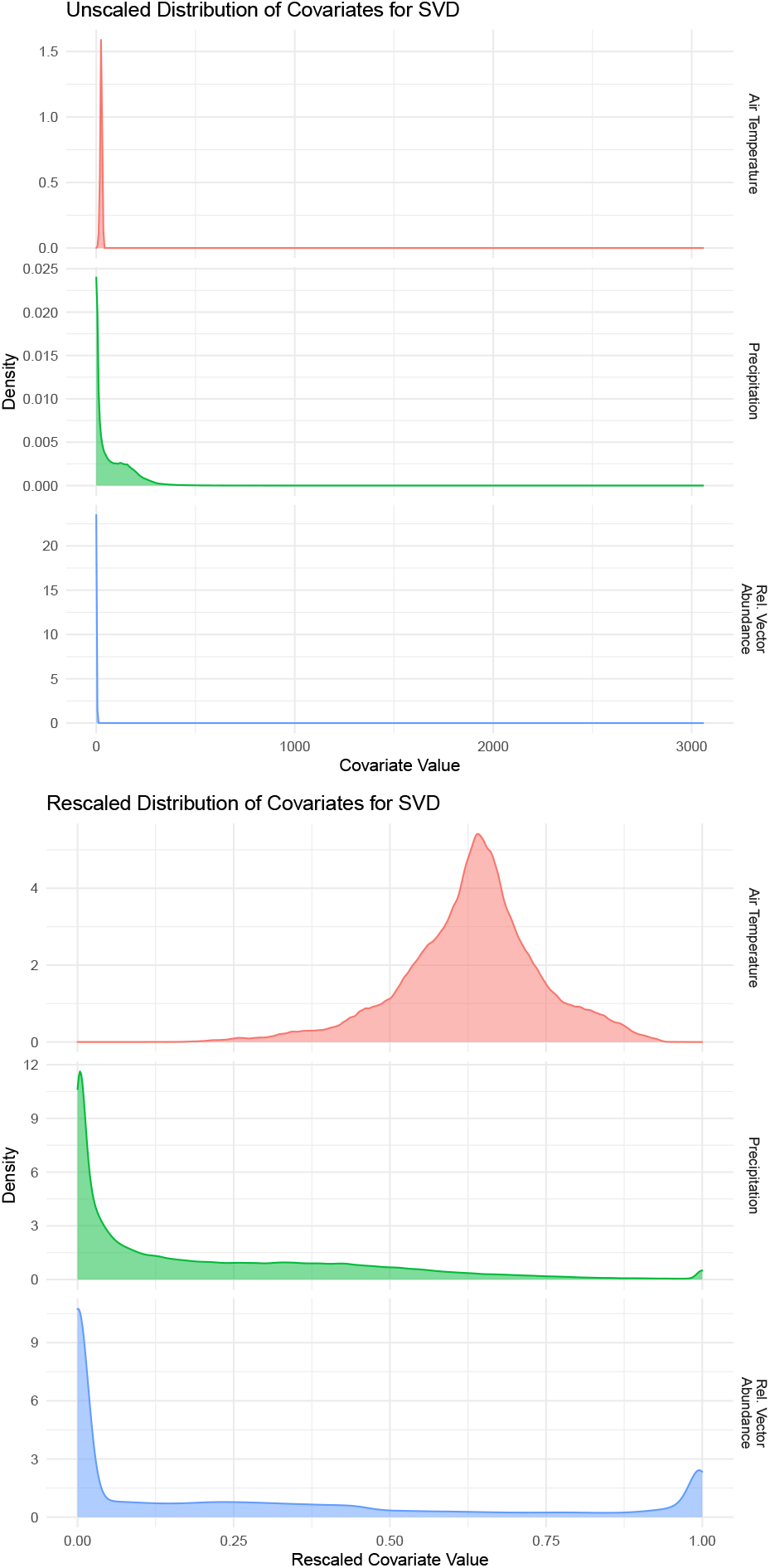
Distribution of covariates for Singular Value Decomposition (SVD) across all pixels before and after rescaling. Synoptic monthly mean temperature (degrees C) and rainfall (mm/month) are from the ERA5 project. Relative vector abundance, a static metric, tracks the proportion of *Anopheles arabiensis, funestus*, and *gambiae* in each pixel.

### Dimensionality Reduction

#### General

If the covariate selection process only locates a few variables of interest, a dimensionality reduction step may not be necessary. However, beyond the point that covariates can comfortably be plotted together (four or five layers at most), dimensionality reduction can be a valuable tool both for data exploration and for more effective clustering. The goal of these techniques is to collapse high-dimensional data into a lower-dimensional space in a way that preserves as much of the original data variation as possible, creating a denser and richer database from a sparser one. A wide range of dimensionality reduction techniques are available in machine learning software packages.

This analysis utilized Singular Value Decomposition (SVD), a strategy similar to Principle Components Analysis which locates the set of orthogonal vectors in a high-dimensional dataset that cover the most variance in the data, and returns those vectors in order of variance explained [24, 25]. SVD is evaluated by plotting how much variance in the original dataset is explained by the most informative singular vectors. If a small number of vectors covers a large portion of the initial variance, then only that smaller-dimensional dataset need be retained for further analysis. While straightforward to implement, SVD is not a time-series method, meaning that dimensionality reduction does not explicitly take into account causal correlations between the different monthly layers of climate data. Other strategies, such as Fourier transforms or dynamic mode decomposition, are worth consideration if simple methods such as SVD do not yield informative results, or if the input data covers long time series rather than synoptic trends.

#### Example Configuration

The final covariate dataset was comprised of 27 dimensions: 12 layers each of rainfall and air temperature data and three layers of relative vector abundance data. SVD was applied using the svd command in R version 3.6.2. Points that are close together in this reprojected space are expected to be similar in terms of their rainfall, temperature, and relative vector proportions. Upon running SVD, the first three singular vectors explained over 95% of the variation in the full dataset, and were retained for clustering (Figure 2).

**Figure 2.**
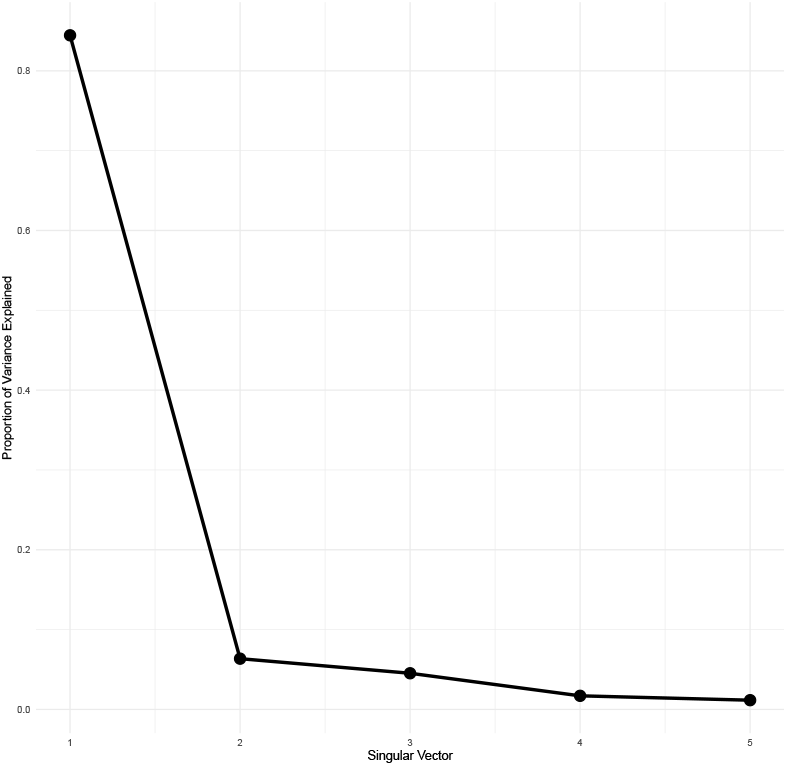
Proportion of variance explained by the first five singular vectors after singular value decomposition (SVD). The first three vectors were retained for clustering analysis.

### Clustering

#### General

The literature and theory on clustering algorithms is vast [26, 27], including many excellent tutorials for beginners. Briefly, clustering strategies are unsupervised learning methods that group data points together based on proximity in all provided dimensions.

The example configuration utilized k-means for clustering. This algorithm benefits from simplicity and relatively quick runtime using an iterative geometric search, but suffers two major setbacks. First, the choice of cluster count *k* must be specified by the user, which can lead to nonintuitive cluster groupings if an inappropriate *k* is selected. Intuitively, the goal of k-means is to minimize data variance within clusters while maximizing it between clusters. A heuristic but common solution for addressing the cluster count problem is the use of “elbow plots”, in which cluster count is plotted against the ratio of between-cluster variance and total variance. This ratio is monotonically increasing, since the cluster count that maximizes this ratio is a *k* equal to the number of data points, but the goal is to find a *k* for which the plot makes an “elbow” and begins to increase more slowly. Other strategies for selecting cluster counts for k-means include the use of Jaccard, Dunn, and Silhouette indices [27].

A second weakness of k-means is its radial search methodology. Because this clustering algorithm minimizes the Euclidean distance between a cluster centroid and the data points around it, clusters that are tightly grouped in all dimensions will be captured more effectively than elliptical or other cluster shapes. Heuristically, this did not appear to affect the algorithm’s ability to find distinct groups, but non-spherical methods such as CLARA or Gaussian Mixture Models (GMMs) may be more appropriate in some settings. GMMs, which utilize a Bayesian fitting procedure, have the added benefit of allowing for the calculation of information criteria, giving a more principled and less heuristic way to select cluster counts compared to k-means.

#### Example Configuration

Having reduced the dimensionality of the dataset from 27 to three, the next step was to classify points into transmission archetypes representing unique environmental and entomological patterns of relevance to malaria transmission. To identify these distinct archetypes, k-means clustering was performed on the reduced-dimensional dataset output by SVD. K-means was run using the MacQueen algorithm [28] on the reduced-dimensional dataset over a range of possible cluster counts from 3 to 15, and elbow plots computed. Unfortunately, these plots did not show a strong elbow (Figure 3), so a cluster count of 10 was selected as a compromise between relatively high variance ratio and a visually intuitive and distinguishable set of archetypes (Figure 4, Figure 5, Figure 6). K-means was applied using the kmeans command in the stats R package version 4.0.5. See the Results section for a more in-depth description of k-means outputs.

**Figure 3.**
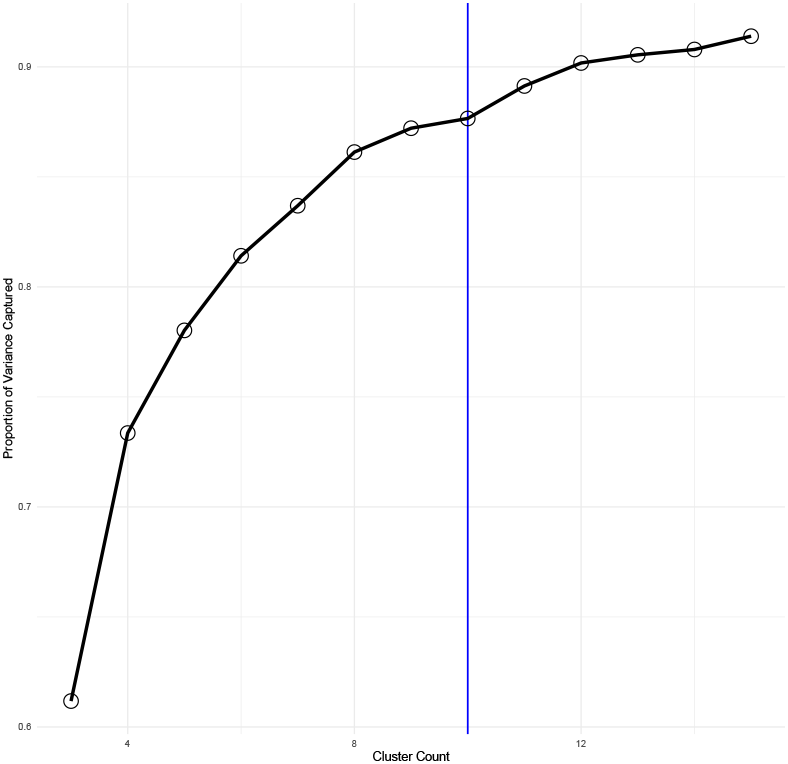
K-means elbow plot for *k* between 3 and 15. The x-axis shows cluster count, while the y-axis shows the proportion of total data variance that is captured by between-cluster variance in each setting. A cluster count of ten (vertical blue line) was chosen for further analysis.

**Figure 4.**
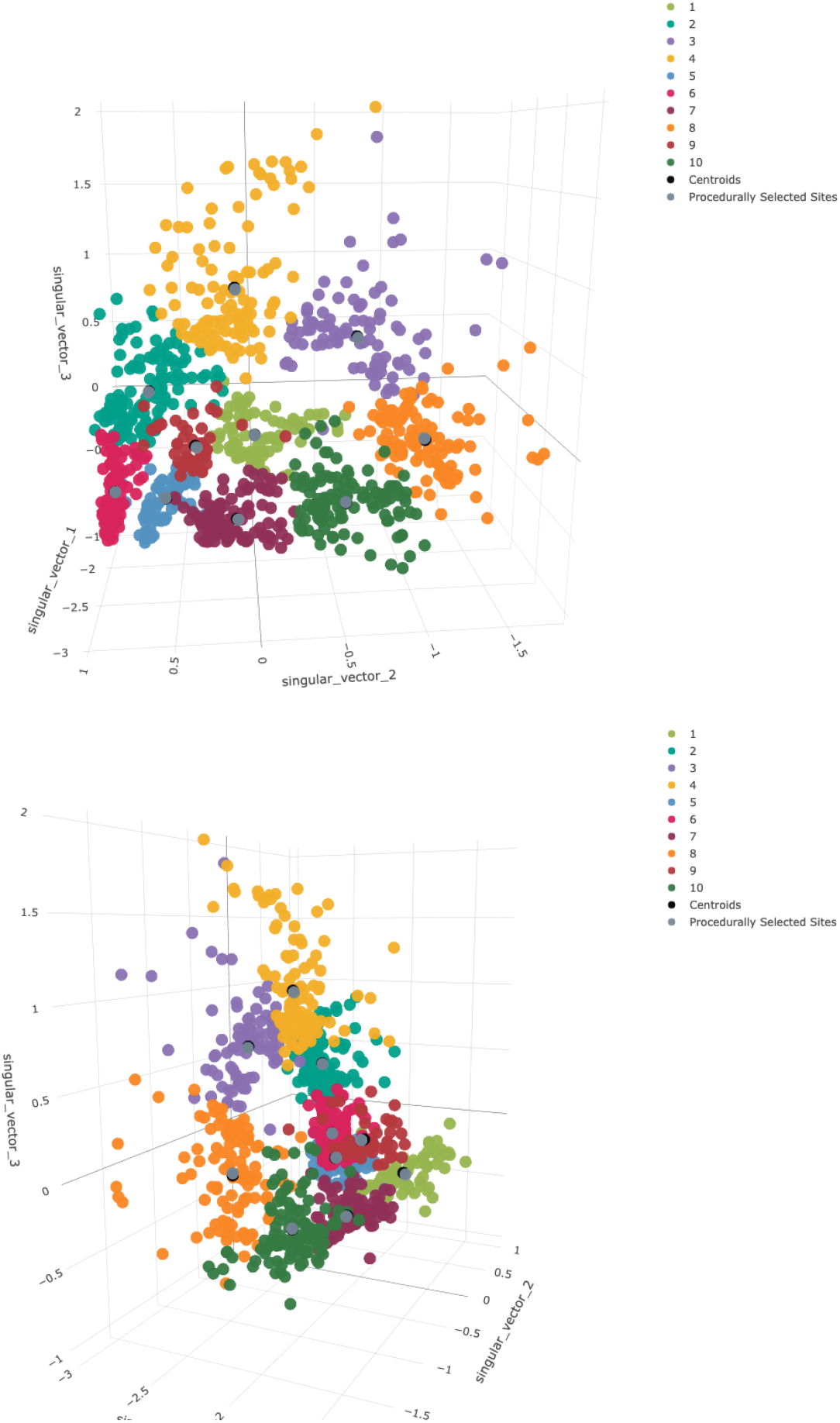
K-means results. Top and side view of pixel values reprojected onto the first three singular vectors, after k-means clustering with a *k* of ten. Colors refer to different clusters and match subsequent six-cluster plots. Black dots indicate true cluster centroids, while dark grey dots show procedurally-selected representative sites.

**Figure 5.**
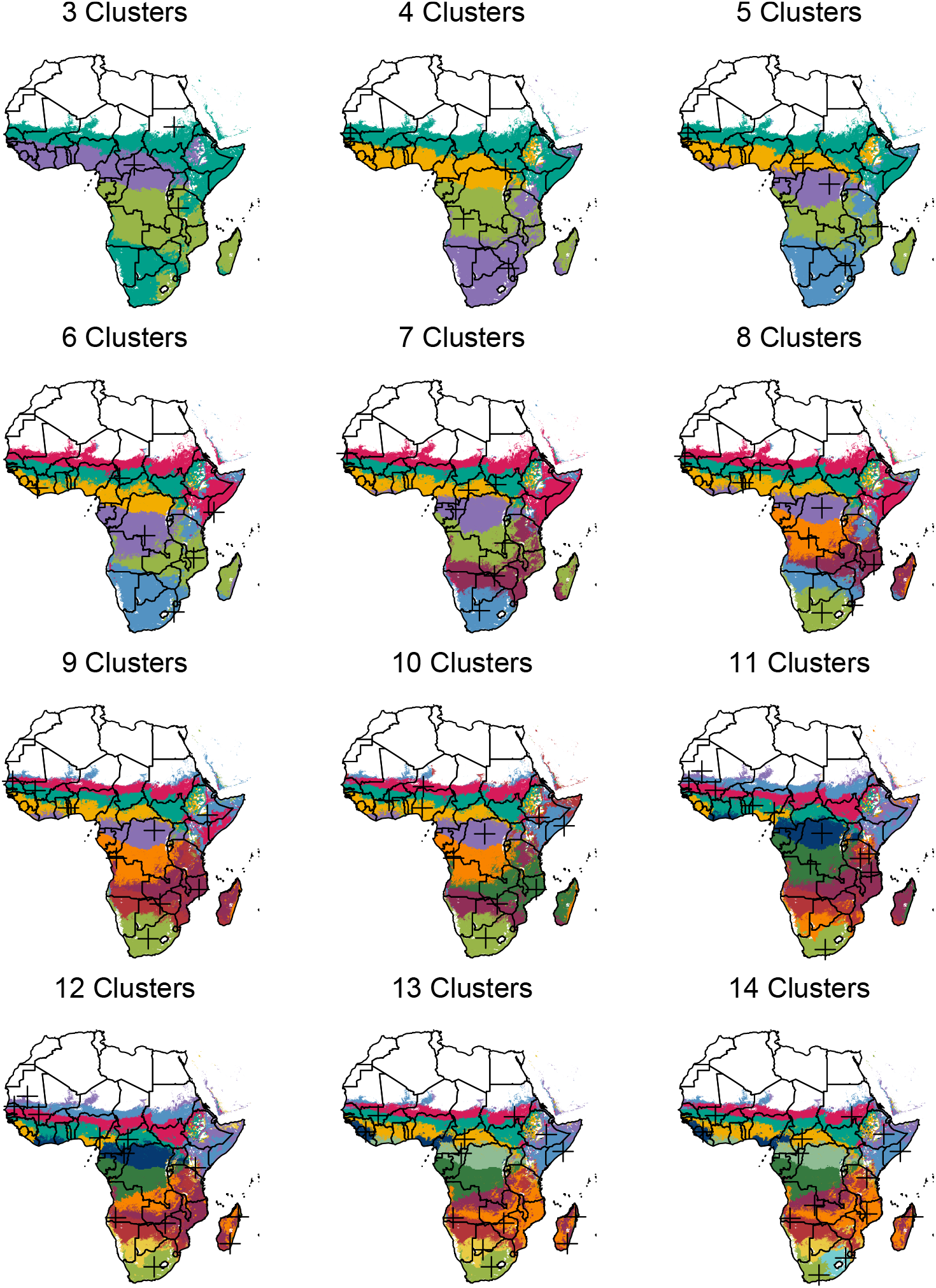
Cluster maps for *k* of 3 to 14. Black crosses indicate procedurally-generated representative sites (the pixels closest to cluster centroids).

**Figure 6.**
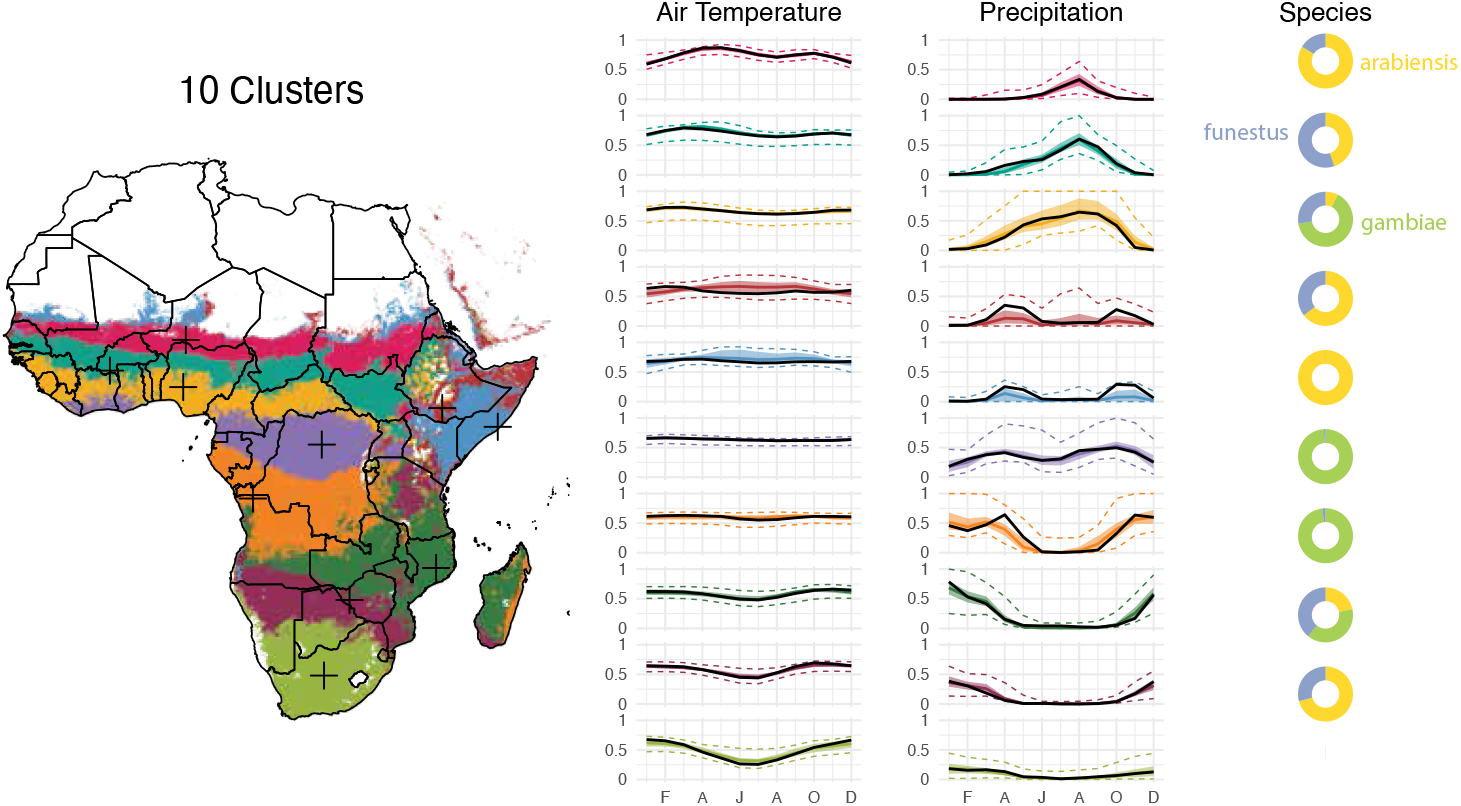
Maps and time series for the ten-site setting. In the time series, solid colored lines represent the median across the archetype, shaded areas indicate the interquartile range, and colored dotted lines indicate the 95% variance interval. Solid black lines represent the procedurally-selected climate values of the representative site for each archetype, also indicated as black crosses on the map. Doughnut plots show the relative vector abundance of the representative sites.

### Selecting Representative Sites

#### General

The clustering step groups the data into archetypes. Next, a set of specific covariate values must be selected for each archetype to use as inputs into the transmission model. One viable option would be to take the mean or median across all data points and use these as model inputs for the archetype, acknowledging that this measure of center will not reflect any true location and may be sensitive to outliers in the clustering process. Using the values at each cluster centroid would be less sensitive to outliers, but still not reflect any true physical location. A third alternative, used in the example configuration, is to select the data point closest to each k-means cluster centroid and use this as a representative site for the archetype as a whole. Visual examination of the k-means clusters (Figure 4) shows that the third approach is nearly identical to the second in the example configuration, as there is generally a trivial distance between the true cluster centroids and their nearest data point. Sensitivity analysis was conducted to assess the acceptability of this approach.

#### Example Configuration

The data point closest to each cluster centroid (determined using k-Nearest Neighbors, with *k* set to one) was selected as the site whose input data would be used for simulation. K-nearest neighbors was applied using the get.knn command in the FNN R package version 1.1.3.

#### Site Selection Sensitivity Analysis

In the main analysis, a single centrally-located point is used as a proxy for all other locations in that cluster. A sensitivity analysis was conducted to assess how other locations in each cluster behaved under the same intervention scenarios. Ten pixels were randomly chosen from each cluster, and all intervention scenarios were re-run on these 1,000 points in the same way that they were on the original ten. Results were compared to those from the cluster centroid sites alone to assess within-cluster variance.

### Selecting and Configuring a Malaria Model

#### General

A number of mechanistic malaria models are commonly used by groups consulting with national or international malaria policy groups. These include OpenMalaria [29], EMOD [14], the Imperial model [18], and VCOM [30]. These models vary with respect to their structure, ease of use, and public accessibility and can produce measurably different estimates under similar initial conditions [31]. Modelers should have a close familiarity with their model of choice prior to embarking on the exercises described in this document. In particular, it is necessary to understand the input parameters to which each model is most sensitive and how this sensitivity impacts covariate and archetype selection; how to appropriately parameterize each site’s demographics, climate, entomology, and interventions; and how to run checks to diagnose unexpected model behavior before acting upon any conclusions from model results.

When pursuing the archetypes strategy, modelers will utilize values from the clustering covariates for some model inputs, but must choose how to initialize all variables not included in the clustering covariates. For instance, in the example configuration, the clustering analysis provided input rainfall, temperature, and entomology variables, but not demographic, intervention history, or transmission intensity variables. As described in detail below, this analysis held most other variables static across all sites, and ran simulations at a range of initial transmission intensities, but different use cases might call for different input parameters in different representative sites.

#### Example Configuration

This modeling analysis was conducted using the EMOD simulation software produced by the Institute for Disease Modeling. See the Appendix for an introduction to EMOD. All simulations were run with an initial population of 2,000 to balance population size with computational constraints, a birth rate of 36.3 live births per 1,000 people per year from the World Bank Database, and a complementary mortality rate to ensure a stable population over time. Each human in the model was assigned a unique risk of being bitten by a mosquito, such that the distribution of biting risk in the community overall was exponential [32].

Simulations were initialized by running one 39-year intervention-free simulation for each of 50 initial transmission intensities and 10 random seeds, generating a collection of 500 baseline populations upon which to test intervention impact. Transmission intensities were varied by scaling mosquito larval habitat capacity, which linearly impacts the number of adult mosquitoes and the level of malaria transmission in the absence of interventions. The intervention-free immunity establishment period should approximate the length of a human life, and is frequently set to 50 years. A 39-year period was selected because this is the duration of time for which climate data was available from ERA5. Ten is a common choice for number of random seeds to test in EMOD, and has been shown to cover the variation in most parameters well. Fifty transmission intensities were chosen to thoroughly cover variability in this important parameter. One hundred fifty-two different intervention packages were tested, covering a wide range of vector control, drug, and vaccine-based malaria prevention strategies. These intervention parameters were originally part of a project to test eradication feasibility in best-case scenarios, and therefore often represent coverage levels or policies more rigorous than those commonly in use today (for example mass bednet campaigns every year instead of every three years). Each intervention was run for three years, and its efficacy assessed by mean *Plasmodium falciparum* parasite rate among 2-10 year-olds (*Pf PR*_2*-*10_) in the final year of the intervention compared to the final year of the intervention-free simulation.

Climate inputs of air temperature, rainfall, and relative humidity were constructed from the publicly-available ERA5 climate reanalysis model. For each representative site, daily climate data from 1980 to 2018 was downloaded from ERA5. The “2m temperature” channel was used for air temperature, the “total precipitation” channel for rainfall, and the “2m temperature” and “2m dewpoint temperature” channels used to calculate relative humidity according to a method developed by the company Vaisala [33]. The intervention-free simulations to establish population immunity were run using climate data from 1980 to 2018, and the three-year intervention simulations were run using climate data from 2016 to 2018.

Mosquito parameters can be found in Table 1, and intervention descriptions and parameters in Table 1 and Table 3.

**Table 1.** EMOD mosquito species parameters.

| Name | Anthropophily (%) | Endophily (%) |
| --- | --- | --- |
| arabiensis | 65 | 50 |
| funestus | 65 | 85 |
| gambiae | 85 | 85 |

**Table 2.**
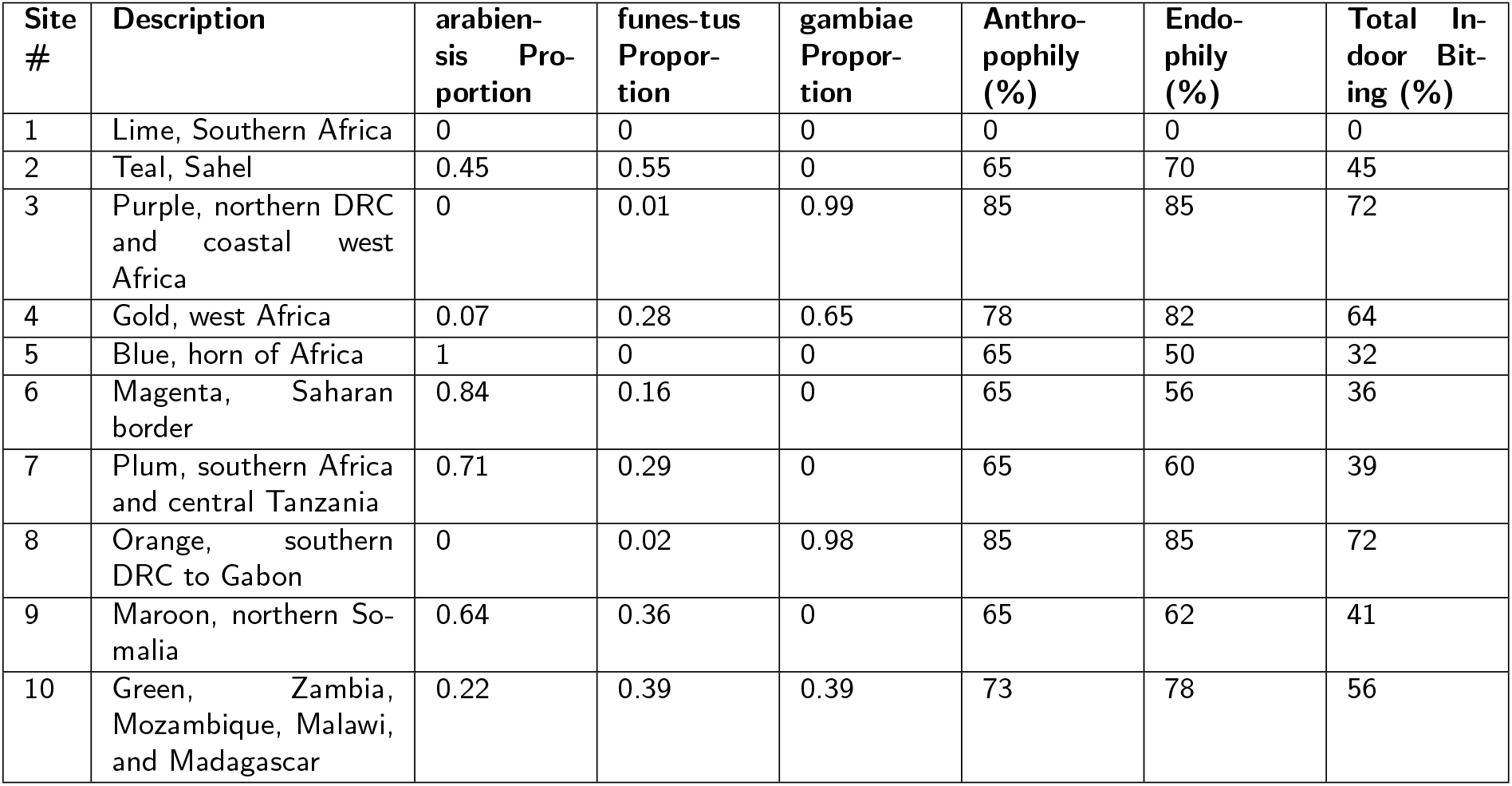
EMOD Site Descriptions.

**Table 3.** EMOD intervention descriptions.

| Intervention | Details |
| --- | --- |
| Insecticide-Treated Nets (ITNs) | Distributed annually on January 1st;<br>Initial blocking 90%, decaying exponentially with a 2-year half-life;<br>Initial killing 60%, decaying exponentially with a 4-year half-life;<br>Nets are discarded with exponential decay and a 6-month half-life;<br>A randomly-selected 10% of the population never receives a net;<br>Everyone who owns a net uses it every night. |
| Indoor Residual Spraying (IRS) | Distributed annually on January 1st;<br>Initial killing 60%, decaying exponentially with a 6-month half-life;<br>No correlation between IRS and ITN coverage. |
| Artemether-Lumefantrine (AL) Case Management | Constant throughout simulation;<br>"Coverage" refers to the probability of seeking care given a clinical case;<br>Upon deciding to seek care, per-day probability of care-seeking is 0.15;<br>No age dependence;<br>Drugs clear infection and provide about two weeks of additional protection. |
| Dihydro-artemisinin-Piperaquine (DP) Case Management | Constant throughout simulation;<br>Same as AL case management, but with 1 month of protection after use. |
| Pre-Erythrocytic Vaccine (PEV) | Constant throughout simulation;<br>90% acquisition blocking, decaying exponentially;<br>6- and 12- month decay half-lives;<br>Distributed to infants upon reaching 6 months of age. |
| Transmission-Blocking Vaccine (TBV) | Distributed annually on January 1st;<br>90% transmission blocking, decaying exponentially;<br>6- and 12- month decay half-lives;<br>Distributed to adults age 15-49. |
| Attractive Targeted Sugar Baits (ATSB) | Distributed twice per year on January 1st and July 1st;<br>Reported in terms of per-feed mortality rate, not coverage;<br>6-month box duration of efficacy. |
| Monoclonal Antibodies (mABs) | Distributed annually on January 1st;<br>Simulated via a PEV (see parameters above);<br>Distributed to adults age 15-49;<br>3-month box duration of protection. |

### Recreating Maps

#### General

Once model simulations have run for each archetype’s representative site, these individual-site results must be reprojected back onto pixel level results. The methodology described below constructs a lookup table to convert from baseline per-pixel transmission to intervention-impacted transmission in each archetype. If transmission intensity was included as a clustering covariate, the lookup table approach would differ in its details but be similar in essence.

#### Example Configuration

Once all EMOD simulations were complete, maps of intervention coverage were reconstructed as follows. First, a pixel-level map of *Plasmodium falciparum* prevalence among children aged 2-10 (*Pf PR*_2*-*10_) was selected from the Malaria Atlas Project. Because these EMOD simulations were initiated from intervention-free scenarios, the MAP estimate of *Pf PR*_2*-*10_ in 2000 was selected as a proxy for malaria prevalence in the absence of any interventions. Then, for each intervention scenario and each pixel *p* in the selected map, the following steps were taken:

1. The archetype *a* to which the pixel *p* belongs was identified;
2. A spline was generated between data points of initial and final *Pf PR*_2*-*10_ across all transmission intensities in archetype *a*;
3. This spline was used to map the initial *Pf PR*_2*-*10_ corresponding to that of pixel *p* and archetype *a* to the final *Pf PR*_2*-*10_ in that intervention scenario;
4. This final *Pf PR*_2*-*10_ was logged as the “intervention impact” of pixel *p*.

This allows for the reconstruction of maps hypothesizing the potential impact of different interventions across the continent.

## Results

Presented here are the results of the example configuration.

### Covariate Scaling, Dimensionality Reduction, and Clustering

Before rescaling, synoptic ERA5 air temperature values ranged from 2.2 to 39.0 degrees Celsius, with a median value of 24.9 degrees and an interquartile range (IQR) from 22.4 to 27.0. ERA5 rainfall ranged from 0 to 3,058 mm/month, with a median value of 40.6 and an IQR from 4.9 to 130.8. Rainfall values were capped at the 99th percentile cutoff of 367 mm. Relative mosquito abundance values ranged from 0 to 1, with continent-wide proportions of 43.0% *arabiensis*, 36.8% *gambiae*, and 20.2% *funestus* among those pixels with mosquitoes. Figure 1 shows covariate distributions before and after rescaling. Both rainfall and mosquito vector abundance show peaks near zero and one, while temperature peaks most strongly in the center of the distribution.

Figure 2 shows the variance explained by the first five singular vectors in the SVD procedure. The first three singular vectors accounted for 95.3% of all variance and were retained for the clustering analysis. Figure 3 shows the elbow plot for the k-means procedure. It shows that, while a *k* of ten captures over 85% of total data variance as between-cluster variance, a slighlty higher cluster count would have covered over 90% of this ratio. Figure 4 shows a snapshot of the reprojected data points in 3D space, colored according to the k-means results. Black dots represent true cluster centroids, while gray dots indicate representative sites. The shape of this data is roughly circular or toroidal, with those sites that have appreciable numbers of mosquitoes and rainfall comprising the edges of the circle and only the low-rainfall, low-mosquito-count lime green vector clustering near the origin. Visual examination shows that the ten-cluster k-means differentiates several distinct groups, but may combine some groups that are visually fairly distinct, such as the two lobes of the turquoise cluster (Cluster 4).

### Archetype Creation and Site Selection

Figure 5 shows archetype maps over a range of cluster counts from 3 to 14. Across all cluster counts, archetypes tend toward geographic contiguity. Given that the clustering covariates themselves demonstrate strong spatial autocorrelation, this result is perhaps unsurprising, but lends algorithmic credence to historical strategies of heuristically grouping regions by climate or seasonality. Also notable is the stability of archetype affiliation across different cluster counts. The three-cluster map identifies a strongly seasonal Sahelian/coastal band across west Africa, a band in central Africa with opposite seasonality to the first, and more arid regions to the north and south. Adding clusters allows separation of the northern and southern arid regions, followed by increasingly distinct latitudinal bands in west Africa and a stable boundary between central Democratic Republic of the Congo (DRC) and northern Zambia and Mozambique. Lake Victoria, Rwanda, and Burundi are also reliably grouped together and distinct from their immediate surroundings, usually matched to an archetype further south. The southern tip of Madagascar is reliably grouped with southern Mozambique, while the rest of the island joins the same archetype as northern Mozambique and Zambia. The eastern coastline of Madagascar sometimes joins the archetype dominated by southern DRC. The ten-cluster map was selected for the example configuration to strike a heuristic balance between variance explained in the elbow plot (Figure 3) and the communicative value of showing archetypes with visually distinct seasonality and mosquito mixes.

Figure 6 shows a map of the ten archetypes used in the example configuration, along with the associated temperature and rainfall time series and relative abundance of different mosquito species. In the time series, solid colored lines represent the median across the archetype, shaded areas indicate the interquartile range, and dotted lines indicate the 95% variance interval. Solid black lines represent the climate values of the representative site for each archetype, also indicated as black crosses on the map. Additional File 1 shows similar maps for all cluster counts tested. The ten-archetype setting captures a detailed range of different climate modalities on the continent. Three latitudinal bands across the Sahara to the coast show seasonal rainfall of increasing magnitude and duration (magenta, turquoise, and gold). Relative *arabiensis* proportions decline, and *gambiae* proportions increase, along the same gradient. These latitudinal bands continue down the western side of the continent, with a bimodal *gambiae*-dominated archetype in northern DRC (purple), a strongly seasonal *gambiae*-dominated archetype in southern DRC (orange), and a more gently seasonal *arabiensis* and *funestus*-dominated archetype across northern Namibia and Botswana into southern Zimbabwe and Mozambique (plum). Archetypes are least contiguous along the continental divide, with Ethiopia, Uganda, and much of western Kenya showing a mix of archetypes. This effect is likely driven by the complex and highly varied topography of the Great Rift Valley, which introduces sharper gradients of temperature, precipitation, and mosquito species than more smoothly-varying landscapes elsewhere on the continent. The horn of Africa is classified into two bimodal archetypes dominated by *arabiensis*, one with a substantial fraction of *funestus* (maroon) and one without (blue). Zambia, northern Mozambique, southern Tanzania, and most of Madagascar comprise a strongly seasonal archetype with a near-even mix of all three vectors (dark green). A broad area of central Tanzania is associated with the lower-rainfall, *arabiensis*-dominated southern band rather than any of the archetypes it borders (plum). The lime green archetype in southern Africa is distinguished by having few to no mosquitoes, and is excluded from simulation analysis. With the exception of the two representative sites in the horn of Africa, which display considerably higher rainfall than their archetypes’ median values, representative site time series line up well with the time series of the archetype medians. These two clusters have fewer members and higher variability than many of the others, which may explain this effect. Table 2 describes the geography and mosquito characteristics of each representative site.

### Simulation Outputs and Intervention Maps

Figure 7 shows example impact curves for each archetype under different intervention scenarios, as well as their translation into maps of predicted impact. The impact curves show pre-intervention malaria prevalence on the x-axis and malaria prevalence after three years of interventions on the y-axis. The line of equivalence, shown in grey, indicates no change between initial and final prevalence, while the colored curves show simulation results across the range of transmission intensities. More effective intervention packages have curves that swing farther toward the lower right corner of the plots.

**Figure 7.**
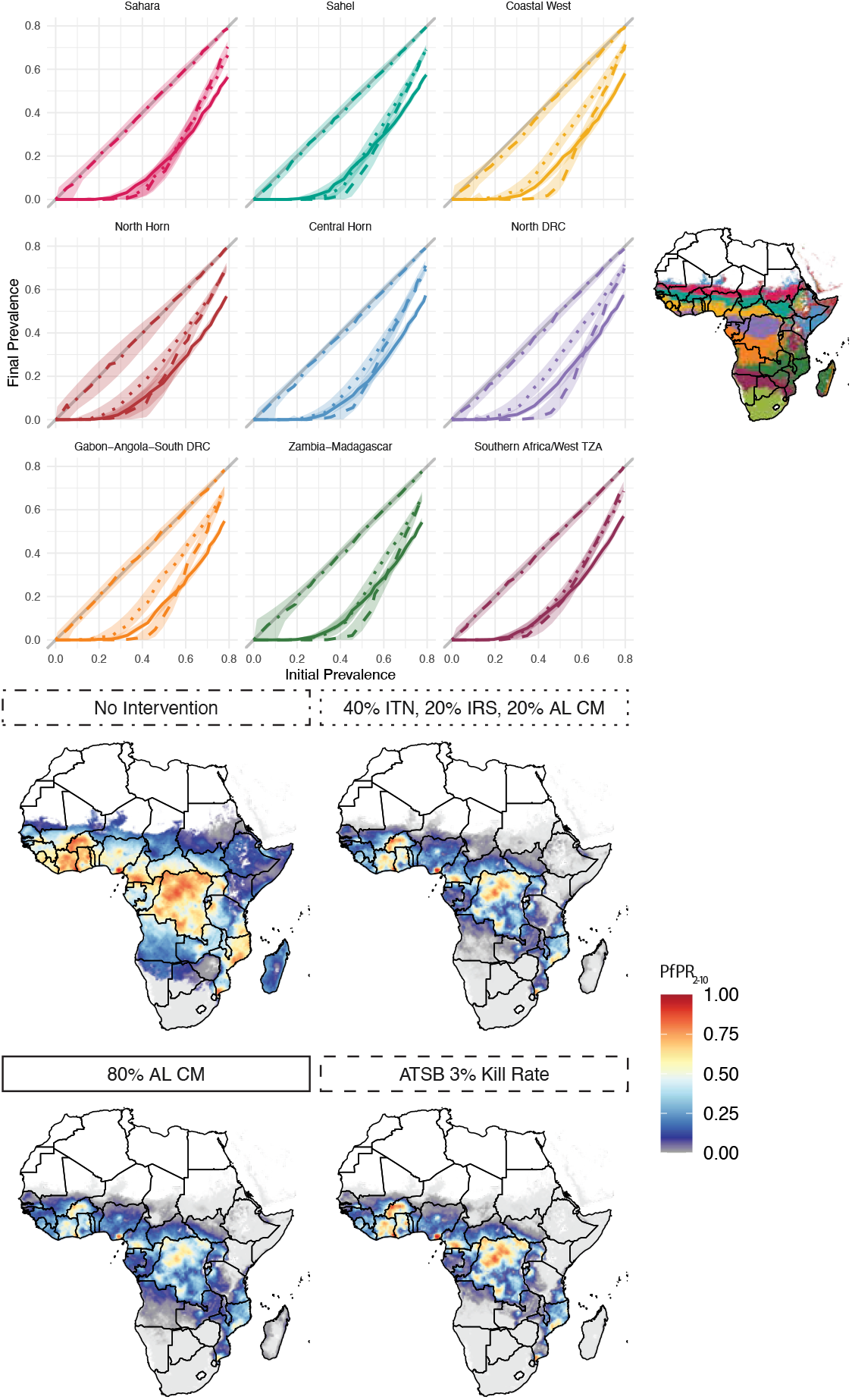
Example intervention impact results for four of the 152 interventions tested. The line plots (top) show intervention impact curves disaggregated by archetype, with archetypes colored according to the map (top right). The four prevalence maps (bottom) show hypothetical *Plasmodium falciparum* parasite rate among children aged 2-10 (*PfPR*_2−10_) for each scenario. These maps were created by starting from a baseline map of 2000 as a proxy for a landscape without malaria interventions. For each initial prevalence value in the baseline map, a new prevalence value was found using the lines in the top set of plots. The linetype of subplot borders in the lower plots matches the linetype of interventions in the upper plots. For more detail about interventions see Table 3. ITN: Insecticide-treated net; IRS: Indoor residual spraying; AL CM: Artemether-Lumefantrine case management; ATSB: Attractive targeted sugar bait.

The first scenario, upper left map and dot-dashed lines, does not have interventions and therefore recreates the baseline 2000 prevalence map. The second, upper right map and dotted lines, shows a complex intervention mix of annual ITN distributions with moderate coverage (40%), annual IRS campaigns with 20% coverage, and low access and use of artemether-lumefantrine case management (AL CM) with, 20% of clinical cases receiving treatment. The third, lower left map and solid lines, shows three years of high antimalarial access and use (80% of clinical cases receiving treatment), but no other interventions. The fourth, lower right map and dashed lines, shows a scenario in which the only intervention is a hypothetical attractive targeted sugar bait (ATSB) with a 3% mortality rate. For more intervention details see Table 3.

In all cases, initial prevalence is the primary driver of intervention impact, but the importance of accounting for archetype is highlighted in comparing scenarios. For example, the upper and lower right maps generally produce similar results despite consisting of non-overlapping interventions. However, difference between the two are clear especially in the plum-colored archetype, in which Angola and southern Zambia have different elimination outcomes between the two exercises and southern Mozambique performs better under an ATSB approach than a complex intervention mix. The antimalarials-only map differs considerably from both of the other intervention packages. In particular, it is more effective than either of the others at high baseline prevalence levels, but often comparable or less effective at low baseline prevalence. This generates a map with a similar elimination landscape, but many fewer hotspots, than the other two. Plots and maps of all 152 interventions are available under the “10-Site Setting” label at https://institutefordiseasemodeling.github.io/archetypes-intervention-impact/.

Each archetype, because of its unique mix of vectors, possesses a different percentage of mosquito bites that occur indoors as opposed to outdoors. It is important for this model framework to capture the differential impact of indoor-biting-targeted interventions in settings with different indoor biting rates. Figure 8 demonstrates this capability. Here, each line represents an archetype, colored according to its indoor biting percentage as determined by its mosquito species mix. The left panel shows an intervention setting with high coverage of insecticide-treated nets and indoor residual spraying, both of which target mosquitoes indoors. This intervention package, as expected, is much more effective in areas with a higher percentage of bites occurring indoors. The right panel demonstrates the inverse property: under an intervention package that includes only anti-malarial drugs and therefore does not directly target mosquitoes at all, there is no differential impact across indoor biting intensities. Table 2 describes the mosquito characteristics of each representative site.

**Figure 8.**
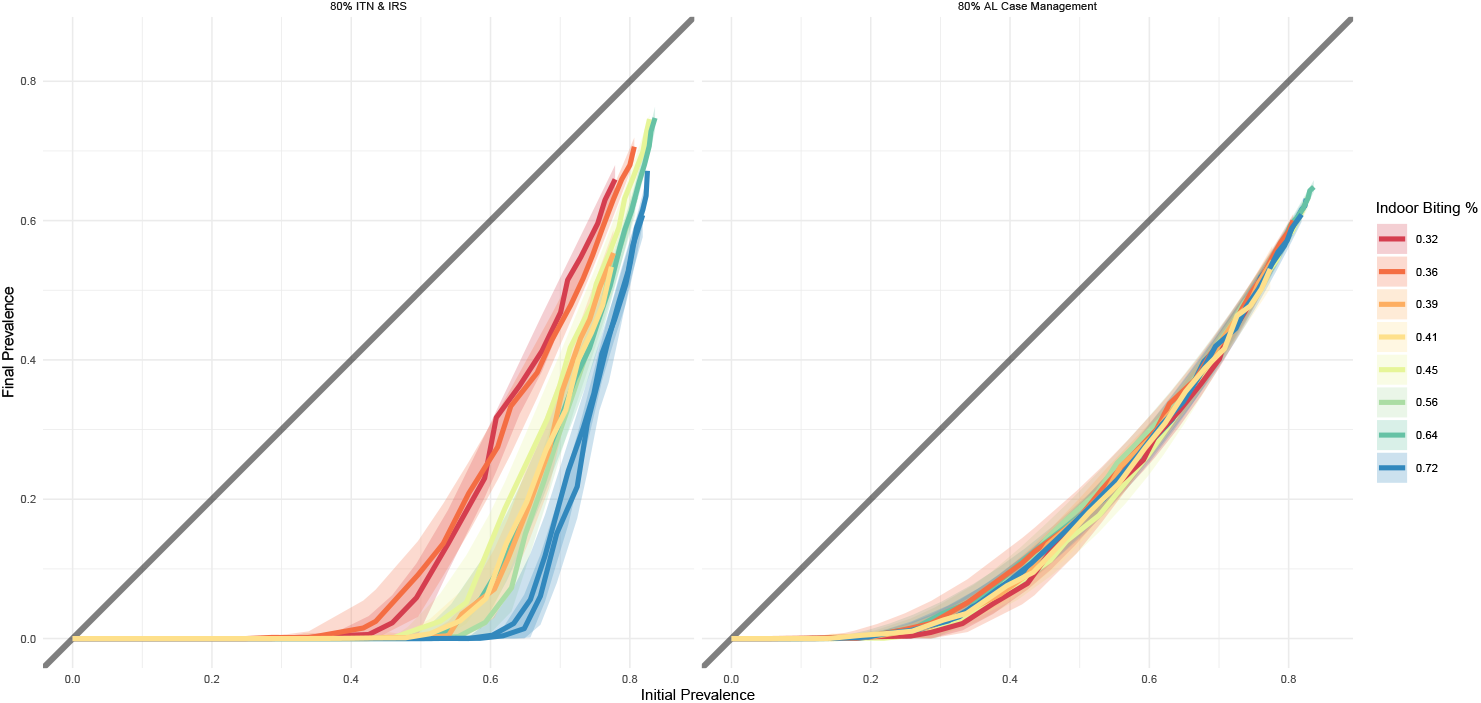
Sensitivity of sites to biting intensity for vector control vs non-vector-control interventions. In both plots, each line represents an archetype’s representative site, colored according to its indoor biting percentage as determined by its mosquito species mix. The left panel shows an intervention setting with 80% coverage of insecticide-treated nets and indoor residual spraying, both of which target mosquitoes indoors. This intervention package, as expected, is much more effective in areas with a higher percentage of bites occurring indoors. The right panel demonstrates the inverse property: under an intervention package that includes only anti-malarial drugs and therefore does not directly target mosquitoes at all, there is no differential impact across indoor biting intensities.

### Site Selection Sensitivity Analysis

Figure 9 shows the location of all 100 sensitivity sites. With one exception, the sensitivity analysis showed that the representative sites appropriately capture variation within archetypes. Figure 10 shows an example intervention package, with the impact curve of each archetype’s representative site plotted in color and the curves of the ten randomly-selected sites shown in black. Shaded areas represent variation across the ten random seeds run for each site. If the representative sites were effective proxies for their archetypes, the colored lines and shaded areas would cover all or most of the black lines and shaded areas. While there are a few archetypes in which the representative sites’ variation is slightly too narrow, especially the yellow and blue curves, in general the representative sites capture both the shape and the variation across the sensitivity sites. The one exception is the plum-colored archetype, in which the representative site curve hews closely to seven of the ten sensitivity sites, but completely misses the curves of the three others. This is perhaps unsurprising since this archetype is the least spatially contiguous, consisting of one band across Namibia, Botswana, Zimbabwe, and Mozambique, and a second vertical swath of southern Kenya and much of Tanzania. The representative site for this archetype is in Zimbabwe, while the sensitivity sites range in location from central Tanzania to deep in the Namibian desert. That all of these sites landed in the same archetype suggest a benefit to including more archetypes in future analyses, or setting constraints on spatial contiguity of archetypes. Similar plots for all 152 intervention packages are available under the “Sensitivity Analysis” label at https://institutefordiseasemodeling.github.io/archetypes-intervention-impact/.

**Figure 9.**
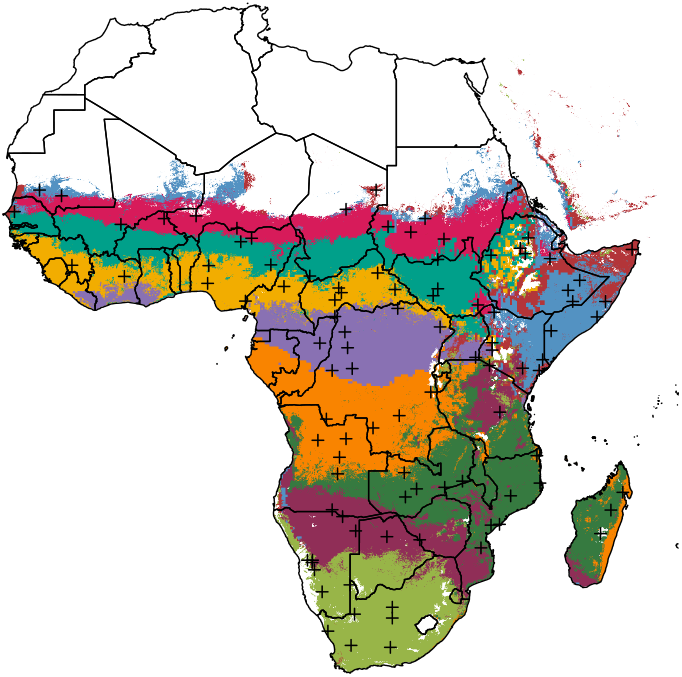
Sensitivity analysis sites. Black crosses indicate the location of all 100 reqpresentative sites used for sensistivity analysis.

**Figure 10.**
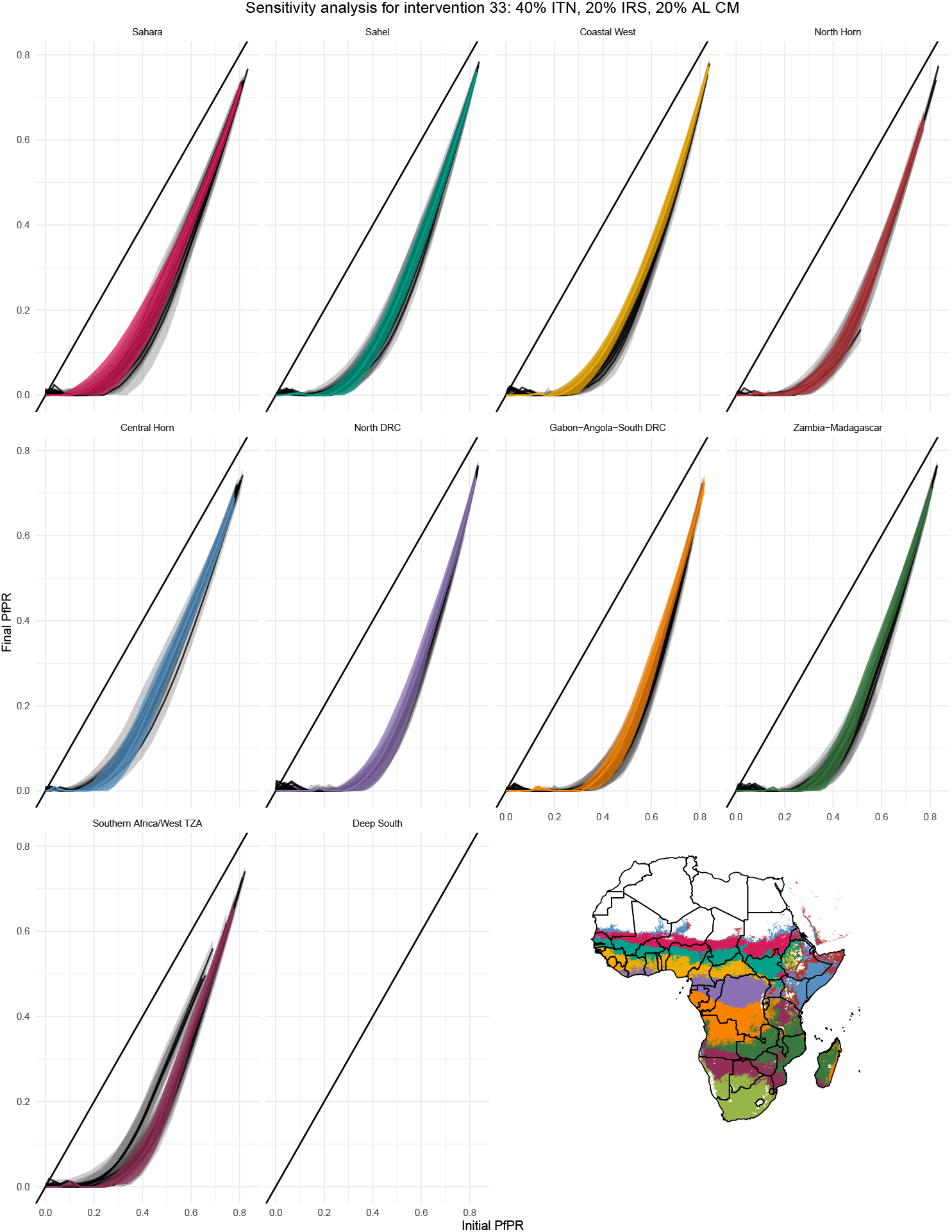
Example sensitivity analysis results. Sensitivity analysis example intervention curves for an intervention package of 40% ITN coverage, 20% IRS coverage, and 20% coverage with artemether-lumefantrine case management. The impact curve of each archetype’s representative site is plotted in color and the curves of the ten randomly-selected sites are shown in black. Shaded areas represent variation across the ten random seeds run for each site. If the representative sites were effective proxies for their archetypes, the colored lines and shaded areas would cover all or most of the black lines and shaded areas.

## Discussion

Mechanistic models are versatile tools for malaria intervention planning, but deploying them over large geographic areas at a useful spatial resolution poses both theoretical and computational challenges. This paper introduces a flexible and customizable archetypes-based framework that harnesses the richness of available spatial data and spatiotemporal modeling products to assess potential intervention impact across a range of settings, and discusses key decision points to consider when constructing such a framework. It includes a detailed example of how the framework might be configured. This example configuration demonstrates that dimensionality reduction and clustering can identify meaningfully different environmental archetypes, that the mechanistic model behaves as expected in relation to intervention effect and mosquito bionomics, that the creation of new maps showing intervention impact can highlight interventions and areas of interest, and that the representative sites selected can appropriately represent each archetype as a whole. The demonstrative analysis used the EMOD mechanistic model and malaria risk surfaces from the Malaria Atlas Project, but this methodology could just as easily be applied with different modeling software or malaria risk estimates. The choice of covariates upon which to cluster is likewise arbitrarily modifiable, noting that the mechanistic model used should have the capacity to accurately reflect any covariate variation that distinguishes archetypes from one another.

An archetypes-based approach can be useful for any project in which there is interest in the differential effects of a disease model under a range of initial conditions. Historically in malaria, such analyses either used idealized “characteristic” settings that did not directly represent real-world locations [18], or tested an extremely wide combinatoric collection of possible initial conditions [11]. The first approach is simple and straightforward, but suffers from arbitrary selection and the inability to claim that the sites shown are actually representative of other areas. The second approach, while thorough, requires an unnecessary computational burden. An archetypes-based strategy provides a useful middle ground, providing a relatively streamlined computational infrastructure for projects that require national or continental coverage, while also offering a useful suite of example settings that can easily be used for any type of exploratory simulation analysis.

The example configuration shows the utility of an archetypes framework for scenario-based analyses that are not calibrated to field data, but the framework has been extended to more data-rich use cases. In the recent High Burden to High Impact (HBHI) initiative, ten high-burden countries developed stratified intervention packages to effectively distribute malaria prevention and treatment to their communities. In Burkina Faso and Nigeria, researchers utilized a covariate-based dimensionality-reduction and clustering strategy to generate subnational archetypes, which then helped inform a rigorous admin-level calibration and stratification process to recommend targeted intervention strategies in upcoming years in these countries [34, 35]. In addition, it has repeatedly proven useful to have a diverse range of archetypal models pre-configured when stakeholders ask policy questions that are not geographically specific but may have different answers in different settings. While identifying different malaria seasonality modalities was not the primary goal of this work, the archetype selection process consistently identified spatial groupings with similar seasonality to heuristically identified seasonality profiles [36, 11]. This observation lends confidence to the effectiveness of both the archetypes framework and other seasonality detection methods.

An archetypes-based approach to intervention impact planning has several limitations. When utilized without fitting to field data, as shown in this document, this method is useful as an intuition-building tool, but should not be used to inform specific decisions in particular geographic locations. While the ability to project results from the analysis to an arbitrarily fine spatial scale may be useful for highlighting heterogeneities, such results might convey an unintended sense of confidence in the sensitivity and specificity of results. It is best to consider this framework as a strategy for obtaining informative priors, rather than as a way of generating quantitiatively rigorous model results. However, as described above in the HBHI framework, these concerns fade when the strategy is utilized as a precursor to more formal model fitting.

The example configuration specifically has additional limitations. While the selected covariates cover many environmental model sensitivities, no human-centered or malaria intervention history covariates were included. While some covariate standardization was performed, a limited number of rescaling or standardizing approaches were conducted. Similarly, while common heuristics were utilized to select singular vector and cluster counts in SVD and k-means, a full and formal sensitivity analysis of these heuristics has yet to be conducted. These would include running k-means with more or fewer singular vectors, and repeating the clustering analysis with k-fold data holdouts. Both of these tests would check how cluster membership changes in response to these varied initial conditions, especially for pixels on the borders between archetypes. However, the robustness of cluster assignment between different k-values provides some confidence that such sensitivity checks would support the choices made in this analysis. While the EMOD modeling software has been extensively vetted and tested, it has not undergone a parameter-by-parameter sensitivity analysis to conclusively determine which variables are most robust to model changes. Because the example configuration focused on hypothetical scenarios rather than prediction or projection, uncertainty was not considered. A full uncertainty propagation strategy would begin with covariate uncertainty and reflect both mechanistic model parameter uncertainty and uncertainty in the prevalence maps used as baseline metrics.

The example configuration additionally does not consider a number of important malaria-related factors, but could easily be extended to take them into account. These include the absence of insecticide resistance, human migration, historical interventions, and site-specific population demographics. Future work will also include model comparison exercises between the archetypes framework and models that have been finely calibrated to specific locations to assess what improvements can be made in archetype-level predictions (while continuing to acknowledge that superseding site-specific analysis is not and will not be the goal). The examples shown here are in Africa, but this framework can also be extended to malaria-endemic regions of Asia and the Americas.

This paper presents a novel archetypes-based strategy for high-resolution, large-area malaria intervention impact assessment. Compared to similar approaches, it adds speed, computational efficiency, and interpretability to results, lends itself well to more detailed calibration-based approaches, and guides intuition in data-sparse settings. [todo: punchy concluding line]

## Data Availability

All data produced in the present study are available upon request to the authors, and will be made available publicly upon publication in a journal.

https://github.com/InstituteforDiseaseModeling/archetypes-intervention-impact

### Appendix

#### Details on the EMOD Software

##### EMOD: An Overview

EMOD (https://idmod.org/documentation) is an individual-based stochastic mechanistic modeling software developed by the Institute for Disease Modeling. While EMOD is capable of modeling a number of infectious diseases, only the malaria framework is described in detail here.

Operating on a daily time step, EMOD simultaneously tracks multiple simulation layers, including but not limited to: mosquito habitat and life cycle, mosquito behavior and movement, human population demographics, human movement, human immune responses to malaria, and malaria interventions that disrupt any of the afore-mentioned layers. Typically, humans are modeled as individuals while mosquitoes are modeled as cohorts, though there are options to model mosquitoes individually for extremely small-scale simulations. As an individual-based model, most input parameters are defined via probability distributions which are sampled at the appropriate points. Spatially distinct communities can be defined, with or without movement between them. While some heterogeneity in risk and behavior can be imposed within communities, under most circumstances a community behaves as a well-mixed population.

##### Mosquito Habitat, Life Cycle, Behavior, and Movement

In EMOD, species-specific mosquito population size is driven by the size of larval habitat available. In the absence of interventions, mosquito abundance in turn drives community-level malaria prevalence. Larval habitat size over time can be specified manually, if entomological data is available to inform this estimate, or can be driven by climate files indicating daily air temperature, rainfall, and relative humidity in the site being modeled. If the climate-driven option is selected, a decision must also be made about the types of larval habitat available in the model, allowing for hydrology-based conversions between climate and mosquito emergence and mortality. For more detail see Eckhoff [14]. Species-specific anthropophily and endophily values can also be specified. In the cohort-based model, mosquitoes do not travel from one community to another. See Table 1 for a list of vector parameters utilized in the example configuration.

##### Human Population Demographics

Humans are modeled individually in EMOD, with each person assigned an age, sex, and “home” community at model initialization. Age distributions and birth and death rates can be specified at model initiation. In addition, individuals can be assigned custom properties related to disease risk or exposure to better capture heterogeneity within communities. For example, in the example configuration, 10% of individuals were assigned to a custom “No Nets” category, to indicate that they would always be skipped during insecticide-treated bednet campaigns, and each individual was assigned a unique biting risk such that the distribution of biting risk in the community overall was exponential [32]. While various types of human movement between communities can be specified in the model, this analysis did not allow for human movement to keep the effects of any one archetypal site clear.

##### Human Immune Responses to Malaria and Accounting for Malaria Population Immunity

Every time a person is infected with malaria in EMOD, they trigger a full intrahost infection cascade. Among the variables tracked are parasitemia, gametocytemia, and var-gene expression. After infection, waning immunity is also tracked and taken into account upon reinfection [37]. Individuals may remain asymptomatic, or develop a mild, clinical, or severe case of malaria. Due to the uncertain pathway from severe malaria to death, malaria-specific mortality was not enabled in this anlaysis.

Upon model initiation, all individuals are immunologically naive to malaria (though a few individuals are initialized as having an active infection). To generate a realistic immune profile, EMOD is typically run with no interventions for several decades, at which point the simulation state can be “frozen”. From this starting point, a range of different intervention scenarios can be initialized.

##### Malaria Interventions

EMOD offers a wide range of malaria interventions, in which the user can specify intervention timing, coverage, targeting, efficacy, and waning, among many other variables. Interventions function by altering default model probabilities– for example, giving an individual an insecticide-treated net would reduce their probability of being bitten by a mosquito indoors, while receiving antimalarial drugs after infection would alter intrahost immune parameters which, in turn, would lower the likelihood of a mosquito acquiring gametocytes and increase the rate of recovery from clinical disease.

Table 3 presents brief descriptions of the malaria interventions used in this analysis. See the EMOD documentation for more.

#### Table of Tested Interventions

**Table 1.** Intervention descriptions and populations at risk (PAR) across sub-Saharan Africa after three years of each intervention package.

| Intervention descriptions and population at risk. |  |  |
| --- | --- | --- |
| Int # | Description | PAR (Millions) |
| 0 | Baseline | 1359 |
| 1 | 0/0/0% ITN/IRS/AL CM | 1319 |
| 2 | 20/0/0% ITN/IRS/AL CM | 1245 |
| 3 | 40/0/0% ITN/IRS/AL CM | 1054 |
| 4 | 60/0/0% ITN/IRS/AL CM | 846 |
| 5 | 80/0/0% ITN/IRS/AL CM | 588 |
| 6 | 0/20/0% ITN/IRS/AL CM | 1105 |
| 7 | 20/20/0% ITN/IRS/AL CM | 965 |
| 8 | 40/20/0% ITN/IRS/AL CM | 722 |
| 9 | 60/20/0% ITN/IRS/AL CM | 548 |
| 10 | 80/20/0% ITN/IRS/AL CM | 370 |
| 11 | 0/40/0% ITN/IRS/AL CM | 789 |
| 12 | 20/40/0% ITN/IRS/AL CM | 617 |
| 13 | 40/40/0% ITN/IRS/AL CM | 455 |
| 14 | 60/40/0% ITN/IRS/AL CM | 335 |
| 15 | 80/40/0% ITN/IRS/AL CM | 236 |
| 16 | 0/60/0% ITN/IRS/AL CM | 484 |
| 17 | 20/60/0% ITN/IRS/AL CM | 357 |
| 18 | 40/60/0% ITN/IRS/AL CM | 274 |
| 19 | 60/60/0% ITN/IRS/AL CM | 195 |
| 20 | 80/60/0% ITN/IRS/AL CM | 158 |
| 21 | 0/80/0% ITN/IRS/AL CM | 261 |
| 22 | 20/80/0% ITN/IRS/AL CM | 209 |
| 23 | 40/80/0% ITN/IRS/AL CM | 164 |
| 24 | 60/80/0% ITN/IRS/AL CM | 125 |
| 25 | 80/80/0% ITN/IRS/AL CM | 106 |
| 26 | 0/0/20% ITN/IRS/AL CM | 1208 |
| 27 | 20/0/20% ITN/IRS/AL CM | 1056 |
| 28 | 40/0/20% ITN/IRS/AL CM | 881 |
| 29 | 60/0/20% ITN/IRS/AL CM | 647 |
| 30 | 80/0/20% ITN/IRS/AL CM | 449 |
| 31 | 0/20/20% ITN/IRS/AL CM | 953 |
| 32 | 20/20/20% ITN/IRS/AL CM | 758 |
| 33 | 40/20/20% ITN/IRS/AL CM | 575 |
| Continued on next page |  |  |

| Intervention descriptions and population at risk, cont. |  |  |
| --- | --- | --- |
| Int # | Description | PAR (Millions) |
| 34 | 60/20/20% ITN/IRS/AL CM | 417 |
| 35 | 80/20/20% ITN/IRS/AL CM | 271 |
| 36 | 0/40/20% ITN/IRS/AL CM | 628 |
| 37 | 20/40/20% ITN/IRS/AL CM | 469 |
| 38 | 40/40/20% ITN/IRS/AL CM | 331 |
| 39 | 60/40/20% ITN/IRS/AL CM | 244 |
| 40 | 80/40/20% ITN/IRS/AL CM | 183 |
| 41 | 0/60/20% ITN/IRS/AL CM | 359 |
| 42 | 20/60/20% ITN/IRS/AL CM | 259 |
| 43 | 40/60/20% ITN/IRS/AL CM | 195 |
| 44 | 60/60/20% ITN/IRS/AL CM | 147 |
| 45 | 80/60/20% ITN/IRS/AL CM | 113 |
| 46 | 0/80/20% ITN/IRS/AL CM | 194 |
| 47 | 20/80/20% ITN/IRS/AL CM | 147 |
| 48 | 40/80/20% ITN/IRS/AL CM | 116 |
| 49 | 60/80/20% ITN/IRS/AL CM | 103 |
| 50 | 80/80/20% ITN/IRS/AL CM | 82 |
| 51 | 0/0/40% ITN/IRS/AL CM | 1053 |
| 52 | 20/0/40% ITN/IRS/AL CM | 870 |
| 53 | 40/0/40% ITN/IRS/AL CM | 674 |
| 54 | 60/0/40% ITN/IRS/AL CM | 490 |
| 55 | 80/0/40% ITN/IRS/AL CM | 331 |
| 56 | 0/20/40% ITN/IRS/AL CM | 760 |
| 57 | 20/20/40% ITN/IRS/AL CM | 557 |
| 58 | 40/20/40% ITN/IRS/AL CM | 403 |
| 59 | 60/20/40% ITN/IRS/AL CM | 289 |
| 60 | 80/20/40% ITN/IRS/AL CM | 204 |
| 61 | 0/40/40% ITN/IRS/AL CM | 441 |
| 62 | 20/40/40% ITN/IRS/AL CM | 321 |
| 63 | 40/40/40% ITN/IRS/AL CM | 236 |
| 64 | 60/40/40% ITN/IRS/AL CM | 168 |
| 65 | 80/40/40% ITN/IRS/AL CM | 122 |
| 66 | 0/60/40% ITN/IRS/AL CM | 252 |
| 67 | 20/60/40% ITN/IRS/AL CM | 186 |
| 68 | 40/60/40% ITN/IRS/AL CM | 138 |
| 69 | 60/60/40% ITN/IRS/AL CM | 112 |
| 70 | 80/60/40% ITN/IRS/AL CM | 90 |
| 71 | 0/80/40% ITN/IRS/AL CM | 137 |
| 72 | 20/80/40% ITN/IRS/AL CM | 112 |
| 73 | 40/80/40% ITN/IRS/AL CM | 93 |
| 74 | 60/80/40% ITN/IRS/AL CM | 74 |
| Continued on next page |  |  |
| Int # | Description | PAR (Millions) |
| 75 | 80/80/40% ITN/IRS/AL CM | 63 |
| 76 | 0/0/60% ITN/IRS/AL CM | 835 |
| 77 | 20/0/60% ITN/IRS/AL CM | 662 |
| 78 | 40/0/60% ITN/IRS/AL CM | 477 |
| 79 | 60/0/60% ITN/IRS/AL CM | 330 |
| 80 | 80/0/60% ITN/IRS/AL CM | 215 |
| 81 | 0/20/60% ITN/IRS/AL CM | 543 |
| 82 | 20/20/60% ITN/IRS/AL CM | 397 |
| 83 | 40/20/60% ITN/IRS/AL CM | 289 |
| 84 | 60/20/60% ITN/IRS/AL CM | 196 |
| 85 | 80/20/60% ITN/IRS/AL CM | 135 |
| 86 | 0/40/60% ITN/IRS/AL CM | 310 |
| 87 | 20/40/60% ITN/IRS/AL CM | 216 |
| 88 | 40/40/60% ITN/IRS/AL CM | 163 |
| 89 | 60/40/60% ITN/IRS/AL CM | 124 |
| 90 | 80/40/60% ITN/IRS/AL CM | 89 |
| 91 | 0/60/60% ITN/IRS/AL CM | 164 |
| 92 | 20/60/60% ITN/IRS/AL CM | 130 |
| 93 | 40/60/60% ITN/IRS/AL CM | 102 |
| 94 | 60/60/60% ITN/IRS/AL CM | 80 |
| 95 | 80/60/60% ITN/IRS/AL CM | 65 |
| 96 | 0/80/60% ITN/IRS/AL CM | 104 |
| 97 | 20/80/60% ITN/IRS/AL CM | 83 |
| 98 | 40/80/60% ITN/IRS/AL CM | 70 |
| 99 | 60/80/60% ITN/IRS/AL CM | 60 |
| 100 | 80/80/60% ITN/IRS/AL CM | 53 |
| 101 | 0/0/80% ITN/IRS/AL CM | 620 |
| 102 | 20/0/80% ITN/IRS/AL CM | 451 |
| 103 | 40/0/80% ITN/IRS/AL CM | 319 |
| 104 | 60/0/80% ITN/IRS/AL CM | 213 |
| 105 | 80/0/80% ITN/IRS/AL CM | 140 |
| 106 | 0/20/80% ITN/IRS/AL CM | 352 |
| 107 | 20/20/80% ITN/IRS/AL CM | 247 |
| 108 | 40/20/80% ITN/IRS/AL CM | 176 |
| 109 | 60/20/80% ITN/IRS/AL CM | 123 |
| 110 | 80/20/80% ITN/IRS/AL CM | 90 |
| 111 | 0/40/80% ITN/IRS/AL CM | 189 |
| 112 | 20/40/80% ITN/IRS/AL CM | 142 |
| 113 | 40/40/80% ITN/IRS/AL CM | 107 |
| 114 | 60/40/80% ITN/IRS/AL CM | 81 |
| 115 | 80/40/80% ITN/IRS/AL CM | 65 |
| Continued on next page |  |  |
| Int # | Description | PAR (Millions) |
| 116 | 0/60/80% ITN/IRS/AL CM | 108 |
| 117 | 20/60/80% ITN/IRS/AL CM | 89 |
| 118 | 40/60/80% ITN/IRS/AL CM | 73 |
| 119 | 60/60/80% ITN/IRS/AL CM | 60 |
| 120 | 80/60/80% ITN/IRS/AL CM | 52 |
| 121 | 0/80/80% ITN/IRS/AL CM | 68 |
| 122 | 20/80/80% ITN/IRS/AL CM | 62 |
| 123 | 40/80/80% ITN/IRS/AL CM | 54 |
| 124 | 60/80/80% ITN/IRS/AL CM | 48 |
| 125 | 80/80/80% ITN/IRS/AL CM | 41 |
| 126 | 80/80/80% ITN/IRS/AL CM + 40% mAb | 38 |
| 127 | 80/80/80% ITN/IRS/AL CM + 40% TBV | 35 |
| 128 | 80/80/80% ITN/IRS/AL CM + 40% PEV | 41 |
| 129 | 80/80/80% ITN/IRS/AL CM + 80% mAb | 37 |
| 130 | 80/80/80% ITN/IRS/AL CM + 80% TBV | 32 |
| 131 | 80/80/80% ITN/IRS/AL CM + 80% PEV | 41 |
| 132 | 80/80/80% ITN/IRS/DP CM | 42 |
| 133 | 0/0/0% ITN/IRS/AL CM + 0.15% ATSB Kill Rate | 1305 |
| 134 | 0/0/0% ITN/IRS/AL CM + 3% ATSB Kill Rate | 423 |
| 135 | 0/0/0% ITN/IRS/AL CM + 15% ATSB Kill Rate | 13 |
| 136 | 0/0/0% ITN/IRS/AL CM + 25% ATSB Kill Rate | 0 |
| 137 | 20/20/20% ITN/IRS/AL CM + 0.15% ATSB Kill Rate | 712 |
| 138 | 20/20/20% ITN/IRS/AL CM + 3% ATSB Kill Rate | 113 |
| 139 | 20/20/20% ITN/IRS/AL CM + 15% ATSB Kill Rate | 9 |
| 140 | 20/20/20% ITN/IRS/AL CM + 25% ATSB Kill Rate | 0 |
| 141 | 40/40/40% ITN/IRS/AL CM + 0.15% ATSB Kill Rate | 206 |
| 142 | 40/40/40% ITN/IRS/AL CM + 3% ATSB Kill Rate | 49 |
| 143 | 40/40/40% ITN/IRS/AL CM + 15% ATSB Kill Rate | 10 |
| 144 | 40/40/40% ITN/IRS/AL CM + 25% ATSB Kill Rate | 0 |
| Continued on next page |  |  |
| Int # | Description | PAR (Millions) |
| 145 | 60/60/60% ITN/IRS/AL CM + 0.15% ATSB Kill Rate | 75 |
| 146 | 60/60/60% ITN/IRS/AL CM + 3% ATSB Kill Rate | 28 |
| 147 | 60/60/60% ITN/IRS/AL CM + 15% ATSB Kill Rate | 0 |
| 148 | 60/60/60% ITN/IRS/AL CM + 25% ATSB Kill Rate | 0 |
| 149 | 80/80/80% ITN/IRS/AL CM + 0.15% ATSB Kill Rate | 39 |
| 150 | 80/80/80% ITN/IRS/AL CM + 3% ATSB Kill Rate | 20 |
| 151 | 80/80/80% ITN/IRS/AL CM + 15% ATSB Kill Rate | 0 |
| 152 | 80/80/80% ITN/IRS/AL CM + 25% ATSB Kill Rate | 0 |

## Acknowledgements

The authors would like to thank all members of the WHO Strategic Advisory Group for Malaria Elimination and the Lancet Commission on Malaria Eradication for their support, commentary, and feedback on this project.

## Funding

This publication is based on research conducted by the Malaria Atlas Project and funded in whole or in part by the Bill & Melinda Gates Foundation, including models and data analysis performed by the Institute for Disease Modeling at the Bill & Melinda Gates Foundation.

This work was supported, in whole or in part, by the Bill & Melinda Gates Foundation (OPP1197730). Under the grant conditions of the Foundation, a Creative Commons Attribution 4.0 Generic License has already been assigned to the Author Accepted Manuscript version that might arise from this submission. This work was also supported by the Telethon Trust, Western Australia.

### Abbreviations

AL CM: Artemether-Lumefantrine case management
ATSB: Attractive targeted sugar bait
DP CM: Dihydroartemisinin-Piperaquine case management
DRC: Democratic Republic of the Congo
GMM: Gaussian Mixture Model
HBHI: High Burden to High Impact
IDM: Institute for Disease Modeling
ITN: Insecticide-treated net
IQR: Interquartile Range
IRS: Indoor residual spraying
MAP: Malaria Atlas Project
mAB: Monoclonal antibody
PEV: Pre-erythrocytic vaccine
*PfPR*_2-10_: *Plasmodium falciparum* parasite rate among children aged 2-10
SVD: Singular Value Decomposition
TBV: Transmission-blocking vaccine
WHO: World Health Organization

## Availability of data and materials

All code, and instructions for analysis replication including data downloads, are available at https://github.com/InstituteforDiseaseModeling/archetypes-intervention-impact.

## Ethics approval and consent to participate

Not applicable.

## Consent for publication

Not applicable.

## Authors’ contributions

ABV, SJB, and PWG conceived the study. ABV, JG, CAB, SJB, and PWG designed the models. ABV wrote and ran all analytical steps. JLP contributed to the design of the machine learning infrastructure. MW and DH wrote a tool to convert ERA5 data into model input files. ABV wrote the first draft of the manuscript. ABV, CAB, JG, JLP, MW, DH, TDH, and PWG interpreted the results, contributed to writing, and approved the final version for submission.

## Additional Files

Additional file 1 — Full Cluster Maps

The plots below show maps and covariate summaries for cluster counts of three to 14. In the time series, solid colored lines represent the median across the archetype, shaded areas indicate the interquartile range, and dotted lines indicate the 95% variance interval. Solid black lines represent the climate values of the representative site for each archetype, also indicated as black crosses on the map. Doughnut plots show the relative vector abundance of the representative sites. Absent doughnuts indicate no mosquitoes in that site.

